# C-reactive protein is not a biomarker of depression severity in drug-naïve obese patients with metabolic syndrome

**DOI:** 10.1101/2025.07.08.25331082

**Authors:** Abbas F. Almulla, Spas Kitov, Tanya Deneva, Maria-Florance Kitova, Lyudmila Kitova, Kristina Stoyanova, Drozdstoy Stoyanov, Michael Maes

## Abstract

**Background:** Metabolic syndrome (MetS) is highly prevalent among adults and is frequently accompanied by depressive symptoms. While high-sensitivity C-reactive protein (hsCRP) has been proposed as a potential indicator of depression, existing evidence remains inconclusive.

**Objective:** This study aimed to determine whether increased serum hsCRP or other immune-metabolic biomarkers are associated with depressive symptoms in drug-naïve individuals with obesity and MetS.

**Methods:** A total of 88 drug-naïve patients with obesity and MetS but without coronary-artery disease were enrolled and serum levels of neuro-immune and metabolic biomarkers were assessed.

**Results:** In MetS, the severity of depression, as assessed using the von Zerssen Depression Rating (VZDR) scale was significantly associated with interleukin (IL)-6, leukocyte numbers, triglyceride x glucose (Tyg) index, low-density lipoprotein cholesterol, Apolipoprotein B (all positively) and mean platelet volume (MPV), visfatin and adiponectin (all negatively). There were no significant associations between hsCRP and severity of depression. In MetS patients, hsCRP is strongly associated with increased leukocyte numbers, alkaline phosphatase, γ-glutamyl transferase, uric acid, platelet numbers and MPV, thereby shaping a distinct subtype of MetS, which is not related to depression.

**Conclusions:** Our findings indicate that depressive symptoms in MetS patients are associated with immune–metabolic biomarkers indicating immune activation, atherogenicity and insulin resistance, but not with hsCRP. The reason is that hsCRP in MetS is a biomarker of a specific MetS subtype that is characterized by megakaryopoiesis, hepatocyte activation, and uric acid production, which were not associated with depression.

## Introduction

Metabolic syndrome (MetS) impacts around 20–25% of the adult population globally and represents a significant public health issue due to its correlation with central obesity, hypertension, dyslipidaemia, insulin resistance, and hyperglycemia (Nolan et al., 2017, Fahed et al., 2022), increasing the likelihood of developing coronary artery disease (Alshammary et al., 2021). Major depressive disorder (MDD) often coexists with MetS, leading to further adverse effects on clinical outcomes and overall quality of life (de Melo et al., 2017). Accumulating studies indicate that MetS may serve as a predictor of depressive symptoms, especially in middle-aged and older individuals (Akbaraly et al., 2009, Akbaraly et al., 2011). However, depressive symptoms may, in turn, induce MetS (Pulkki-Råback et al., 2009, Gurka et al., 2016).

Depressive symptoms in various conditions are linked to changes in lipid profiles, insulin resistance, hypertension, and indicators of hepatic dysfunction— pathophysiological characteristics frequently seen in individuals with MetS (Akbaraly et al., 2009, Zelber-Sagi et al., 2013, de Melo et al., 2017, Villarreal-Zegarra and Bernabe-Ortiz, 2020, Maes et al., 2023). MetS and mood disorders exhibit shared immuno-inflammatory and oxidative stress pathways, characterized by elevated atherogenic indices and diminished levels of endogenous antioxidants and anti-inflammatory mediators (de Melo et al., 2017).

Elevated high-sensitivity C-reactive protein (hsCRP), a well-established marker of systemic inflammation, has been consistently observed in individuals with MetS (Manoj Sigdel, 2014, Shih et al., 2022). Increased hsCRP levels are associated with a heightened risk of depressive symptoms including in people with metabolic or cardiovascular comorbidities (Heisey et al., 2024, Ji et al., 2024). Recent findings propose to stratify depressed patients based on their hsCRP levels as “inflammatory depression” (if hsCRP ≥3 mg/L) (Wessa et al., 2024). However, some authors observed that the slight association between hsCRP and depressive symptoms disappeared after adjusting for body mass index (BMI), indicating that obesity may mediate or confound this relationship (Douglas et al., 2004, Khan et al., 2020). Other (Tully et al., 2015, Ji et al., 2024), but not all (Krogh et al., 2014b) studies indicate a positive correlation between hsCRP levels and depressive symptoms. Furthermore, low serum levels of hsCRP demonstrate a lack of specificity as a biomarker for depression, given their overlap with indicators of MetS, subclinical atherosclerosis, and increased BMI (Maes, 2025).

Adipokines have garnered attention for their role in linking metabolic and mood disorders. Elevated leptin, primarily produced by adipocytes, has been consistently associated with an increase in depressive symptoms, while adiponectin typically exhibits a negative correlation (Labad et al., 2012, Chirinos et al., 2013). Moreover, leptin levels exhibit a positive correlation with hsCRP, while adiponectin levels show a negative correlation (Shamsuzzaman et al., 2004, Komatsu et al., 2007). This underscores a significant relationship between systemic inflammation and metabolic dysregulation. The intricate relationship between MetS and depressive disorders, especially via neuroimmune mechanisms, necessitates that research on immune and metabolic profiles in MDD considers stratification according to the presence of MetS (Maes et al., 2024a). Nevertheless, the associations between depression on hsCRP and other immune-metabolic biomarkers in drug-naïve obese patients with MetS and without coronary artery disease (CAD) have remained elusive.

Hence, this study investigates the association between depressive symptoms and hsCRP and other immune-metabolic biomarkers in patients with drug naïve MetS without CAD. The immune-metabolic biomarkers encompass lipid profiles, apolipoproteins A1 (ApoA1) and ApoB, atherogenic indices, insulin resistance biomarkers, adipokines (leptin, adiponectin, visfatin), inflammatory cytokines (TNF-α, IL-6, hsCRP), blood cell indices such as white blood cell count, platelet count, mean platelet volume (MPV), serum uric acid, and liver enzymes, i.e,, gamma-glutamyl transferase (γ-GT) and alkaline phosphatase (ALP). Based on the current state-of-the art knowledge, the specific hypothesis is that depressive symptoms in the target population are more closely linked to underlying metabolic abnormalities than to hsCRP levels alone.

## Participants and Methods

### Participants

Figure 1 shows the flow of the patient through the study. 302 obese patients with BMI over 30 kg/m2, waist circumference over 80 cm for women and 94 cm for men, aged 35-55 years and previously untreated, were screened. All underwent clinical and laboratory screening to select participants with MetS. The diagnosis was made according to the recommendations of the International Diabetes Federation (Alberti et al., 2009) as the presence of three or more of the following criteria: waist circumference (according to the guidelines set by the International Diabetes Federation for Europeans): waist circumference over 80 cm for women and 94 cm for men, body mass index over 30, triglycerides ≥1.7 mmol/l; HDL cholesterol < 1.0 mmol/l in women or < 0.9 mmo9l/l in men, blood pressure ≥130/85 mmHg or those taking medication for hypertension, fasting blood sugar ≥5.6 mmol/l or history of taking antidiabetic medication. Ischemic heart disease was excluded in all patients with exercise stress testing, CT angiography, or selective coronary angiography according to European Guidelines (Vrints et al., 2024).

**Figure 1.**
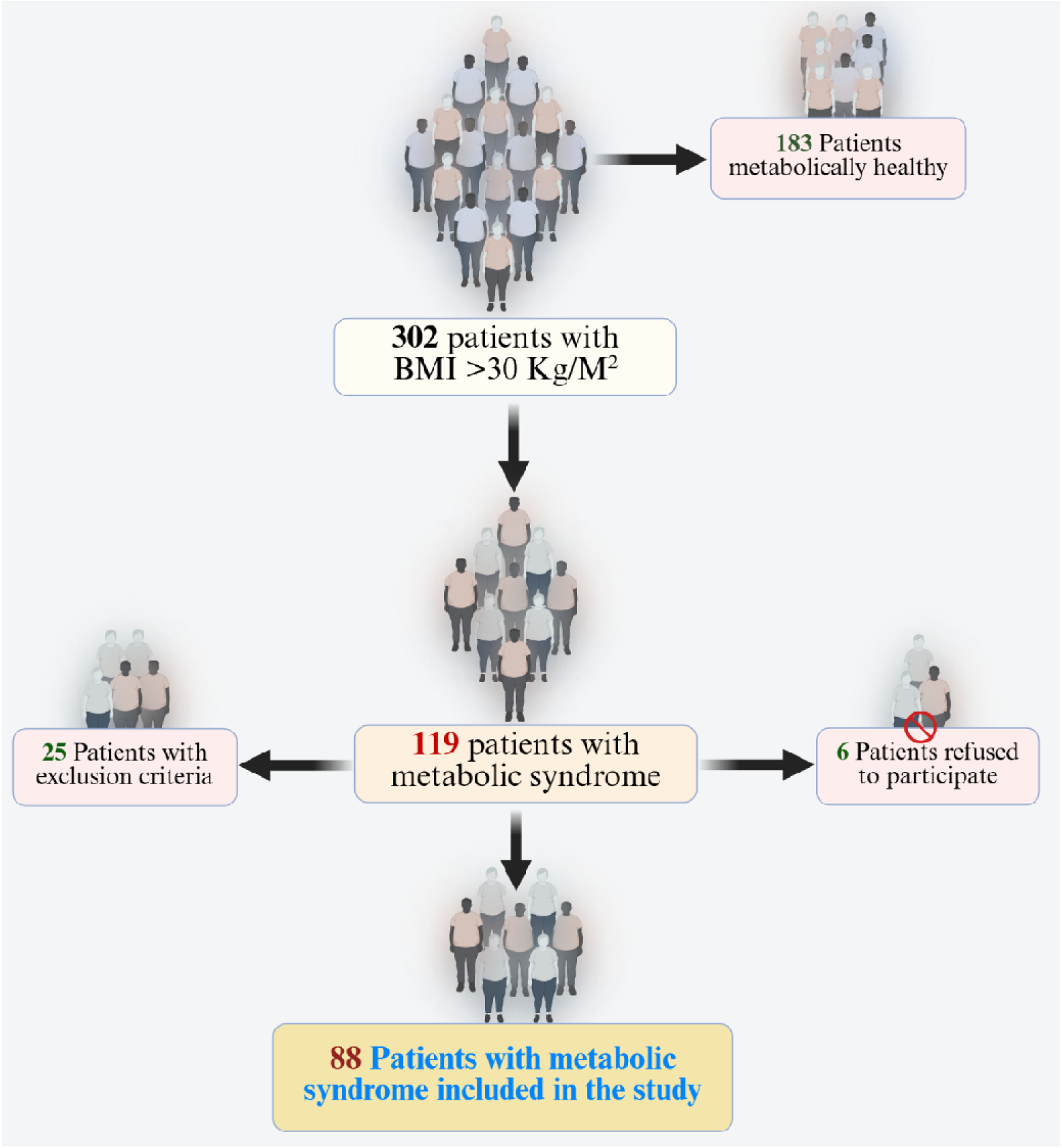
Patient selection algorithm.

Other exclusion criteria were: chronic renal failure, chronic liver disorders, chronic lung disease, moderate or severe valvular heart disease, congenital heart disease, left ventricular systolic dysfunction on echocardiography, pregnancy, known malignancy; thyroid disease, electrolyte imbalance, conduction disorders, as well as patients taking medications known to affect the ECG repolarization parameters, and patients who have had an infection less than 2 weeks ago. We also excluded subjects with neuroinflammatory or neurodegenerative disease, immune disorders, including inflammatory bowel disease, COPD and rheumatoid arthritis, and those with lifetime major psychiatric disorders. During the selection of suitable patients, 25 patients were excluded due to concomitant CHD or refusal to participate in the study, and 6 refused to participate. Finally, a group of 88 patients with newly diagnosed metabolic syndrome aged 35-55 years and without concomitant CHD was recruited.

### Clinical assessments Adiposity-related markers

Using the InBody_270_ analyser (InBody Co, Ltd, Korea) we assessed anthropometric measurements including body weight, height, body mass index (BMI, i.e., weight in kilograms divided by height in meters squared), waist-to-hip ratio (WHR), total body fat percentage. Waist circumference was measured at the level of the umbilicus. Waist-to-hip ratio was calculated from waist circumference measured at the midway between the last palpable rib and the iliac crest and hip circumference (World Health, 2011). Body fat percent was estimated based on multifrequency bioelectrical impedance analysis. The latter technique uses varying frequencies of alternating current that are sent through the body, where passing through different tissues alters the resulting voltage, which information is later used to predict BF%. This renders reasonable estimates compared with the reference method dual-energy X-Ray absorptiometry (McLester et al., 2020). Blood pressure was measured early in the morning, after 10 minutes of rest in a quiet room, while subjects were in a sitting position and using a validated oscillometric device with an appropriately sized cuff applied to the right upper arm. The average of three blood pressure readings (obtained at 1-minute intervals) was taken as the final result.

To assess severity of depression, we used the von Zerssen Depression Rating (VZDR) Scale (Krastev, 2020), translated into Bulgarian. Nevertheless, using principal component analysis, it appeared that no general factor could be extracted from the 16 scale item scores. In fact, the scree plot showed that 5 PCs could be extracted with eigenvalues > 1 explaining only 58.8% of the variance. As such, the total sum on this rating scale does not reflect the total severity of illness. We found that one PC could be extracted from 6 items, which explained 52.2% of the variance, whereby all 6 items showed higher loadings (>0.7). These items are: “I am afraid of losing my mind”, “I feel melancholic and depressed”, “I would like to take my life”, “I often feel simply miserable”, “I cannot think straight”, and “I don’t have any feelings anymore”. These items are indeed indicative of a more severe depression phenotype. Therefore, we used the sum on those 6 items as an index of severity of depression (labeled as VZDR6) together with the total sum (VZDRtotal).

### Blood biomarker assays

Fasting venous blood samples were collected in accordance with standard procedures and serum from each participant was aliquoted and stored at −20°C until analysis, with a maximum storage time of two months.

**Table 1** presents the reference values of the indicators tested in the Central Institute Clinical Laboratory of MU-Plovdiv, Plovdiv, Bulgaria. Hematological analyses, i.e., number of leukocytes and platelets, mean platelet volume (MPV) were performed using the automatic hematological analyzer Advia 2120i using the original reagents from the manufacturer (Siemens Healthcare, Germany). The intra-assay coefficient of variation (CV) for all analytes was between 0.67 and 2.96%, and the inter-assay CV values were between 0.93 to 3.39%.

Biochemical parameters (blood glucose, LDL-cholesterol, total cholesterol, HDL-cholesterol, triglycerides, Apo A1, Apo B, uric acid, HbA1c, alkaline phosphatase, γ-GT were determined using standardized methods recommended by the International Federation of Clinical Chemistry and Laboratory Medicine (IFCC). The tests were carried out using Beckman Coulter reagents on an automated clinical chemistry analyzer Olympus AU 480 (Beckman Coulter, Inc., USA) according to original programs. The analytical intra-assay reliability for all indicators was between 0.37 and 1.06%, and the inter-assay CV was between 0.67 and 2.19%. Serum hsCRP levels were measured by immunoturbidimetry with latex agglutination using an AU 480 Beckman Coulter clinical chemistry analyzer (Olympus AU 480, Beckman Coulter, Inc., USA). The intra-assay and inter-assay CV values of this assay were < 4.87%. Serum insulin was assayed using a one-step immunoenzymatic sandwich method based on a chemiluminescent principle c (CLIA) on the Access 2 Immunoassay System (Beckman Coulter, Inc, USA). The intra-assay and inter-assay CV values of the insulin assay were < 4.2%. The serum concentrations of TNF-α, IL-6, leptin (LDN, Germany), visfatin, and high sensitivity adiponectin were determined using competitive enzyme-linked immunosorbent assays (BioVendor, USA), following validation at the local level. The methods show high precision with intra-assay CV < 10% and inter-assay CV < 12%.

Consequently, we computed 6 different indices, namely Castell risk index 1 as total cholesterol / HDL and the ApoB / ApoA1 ratio, which are both pro-atherogenic indices; LDL / ApoB or LAR index, reflecting the size and composition of LDL particles with smaller particles being more pro-atherogenic; the HOMA-IR index, which estimates insulin resistance; the TyG index = Ln [fasting triglycerides (mg/dL) x fasting blood glucose (mg/dL) / 2), which is a surrogate marker of insulin resistance; and the adiponectin / leptin ratio as a marker of dysfunctional adipose tissue (Frühbeck et al., 2019).

### Data analysis

In the current study we employed IBM SPSS Statistics for Windows Version 30 to conduct all statistical analyses. Differences in continuous variables across groups were assessed using analysis of variance (ANOVA), while analysis of contingency tables using the chi-square (χ²) test was utilized to investigate relationships among categorical variables. Bivariate relationships among continuous variables were examined utilizing Pearson’s product-moment correlation coefficients. Multiple comparisons were adjusted for False-Discovery Rate (FDR) p-correction. Multivariate regression analyses were performed to identify predictors of the severity of depression. A manual and stepwise approach was employed to determine the most significant explanatory variables and generate partial regression plots. Key assumptions, including multivariate normality, homoscedasticity, and the absence of collinearity and multicollinearity, were verified prior to constructing the final regression models. A significant threshold of p < 0.05 was utilized for all statistical tests, employing a two-tailed methodology.

Principal component analysis (PCA) was employed to extract latent constructs from the clinical items and biomarker sets. The Kaiser–Meyer–Olkin (KMO) statistic was employed to assess sampling adequacy, with values exceeding 0.7 indicating sufficient factorability. A principal component is considered valid when the explained variance surpasses 50%, Cronbach’s alpha for internal consistency exceeds 0.70, and all factor loadings on the first component are greater than 0.7. A two-step cluster analysis was employed to generate clusters of patients based on biomarker values. The silhouette coefficient of cohesion and separation was used to assess the quality of the cluster solution, with values exceeding 0.5 indicating adequate cohesion and separation.

The primary statistical analysis is the multiple regression analysis with severity of depression scores as outcome variables and up to 5 covariates. Power analysis showed that when using an effect size of 0.25, p=0.05, power=0.8, and max 5 predictors, the minimum number of subjects should be n=58.

## Results

### Socio-demographic and clinical characteristics of the study group

Figure 2 shows the bimodal data distribution of the hsCRP data. Visual binning revealed that a hsCRP value > 13.5 mg/L could be used to dichotomize the patient group into 2 subgroups, namely those with low (n=50) versus high (n-38) hsCRP levels. **Table 2** compares the socio-demographic and clinical variables among MetS patients categorized by hsCRP levels, utilizing a cutoff of 13.5 mg/L. No statistically significant differences were found between the both high-hsCRP and low-hsCRP groups for all variables assessed.

**Figure 2.**
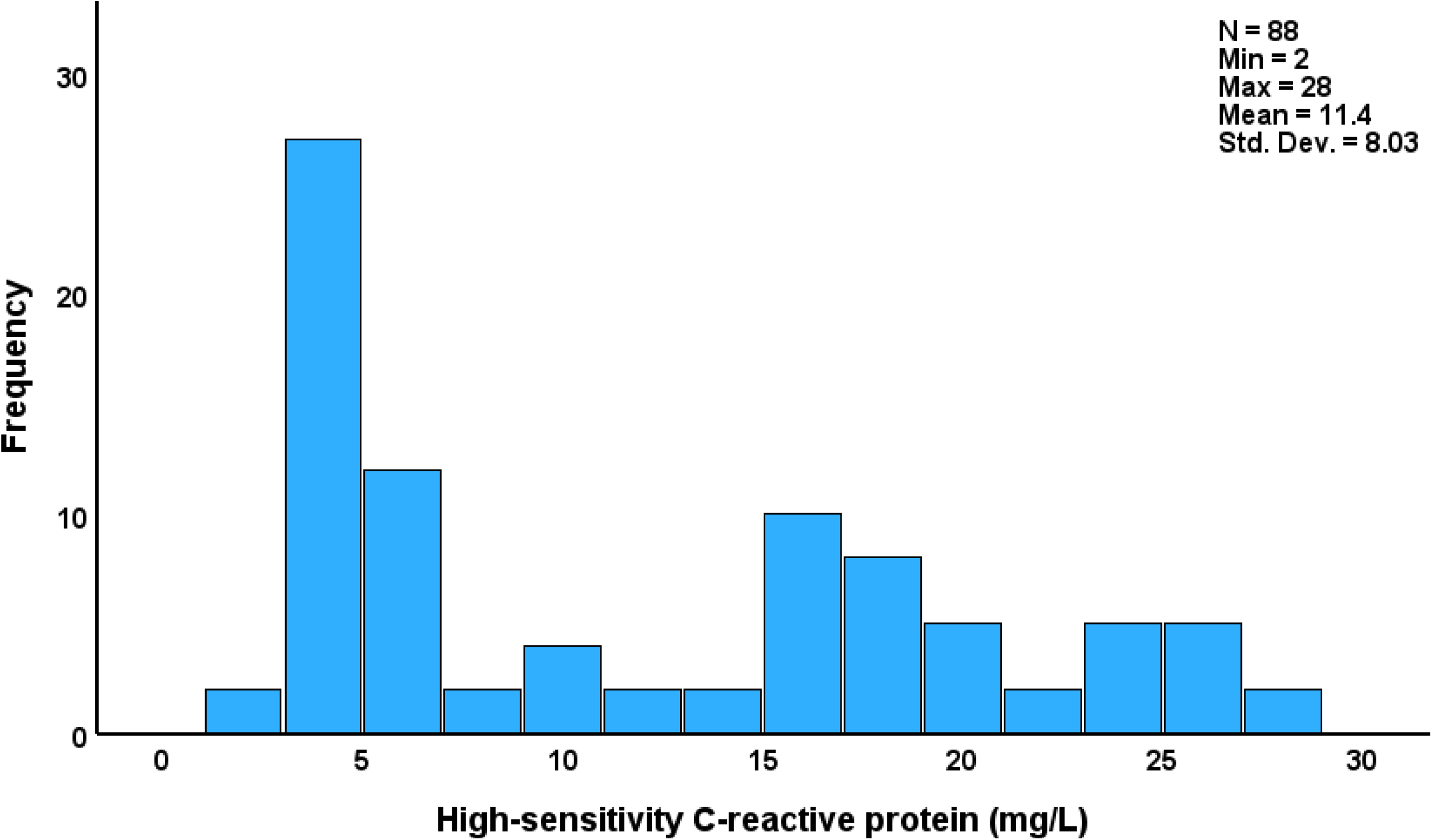
Distribution of high sensitivity C-reactive protein serum levels in patients with metabolic syndrome and obesity.

**Table 1.** The biomarkers examined in the study and their reference values.

| Biomarkers | Units | Reference range in our laboratory |
| --- | --- | --- |
| **1. Immune biomarkers** |  |  |
| Interleukin (IL)-6 | pg/ml | < 17 |
| Tumor necrosis factor (TNF)-α | pg/ml | <12.4 |
| High sensitivity C-reactive protein (hsCRP) | mg/L | < 3: normal<br>>3-10: normal or slightly elevated (obesity, MetS, minor infection)<br>>10-100: moderate inflammation, immune disorders<br>>100: acute bacterial and viral infections |
| **2. Hematology** |  |  |
| Number leukocytes | $10^9/l$ | 3.5-10.5 |
| Number platelets | $10^9/l$ | 140-400 |
| Mean platelet volume (MPV) | fL | 7.5-11.5 |
| **3. Insulin resistance and atherogenicity biomarkers** |  |  |
| Fasting blood glucose (FBG) | mmol/l | 2.8-6.1 |
| Immunoreactive insulin | mIU/l | 1.9-23 |
| HOMA index = FBG x insulin/22.4 |  | < 2.5 |
| Glycated hemoglobin (HbA1c) | % | 4.5-6.3 |
| Total cholesterol (TC) | mmol/l | 3.0-5.2 |
| Triglycerides (TG) | mmol/l | 0.4-1.7 |
| High-density lipoprotein-cholesterol (HDL-C) | mmol/l | >0.9 male<br>>1.15 female |
| LDL-cholesterol | mmol/l | < 1.8 mmol/l high CV risk<br>< 2.6 mmol/l moderate CV risk |
| TC/HDL-C ratio (Castelli risk index1) | - | ≥ 5: increased risk |
| TyG index = $TG \times FBG / 2$ | | 4-8 |
| Apolipoprotein-B (ApoB) | g/l | 0.63-1.88 male<br>0.56-1.82 female |
| Apolipoprotein-A1 (ApoA1) | g/l | 1.09-1.84 male<br>1.06-2.28 female |
| **4. Adipokines** |  |  |
| Adiponectin | mg/mL | 5-30 |
| Leptin | ng/ml | 3,7-11,1 female<br>2,0-5,6 male |
| Visfatin | ng/mL | 0.2 - 1.5 |
| **5. Biochemistry** |  |  |
| Uric acid | $\mu\text{mol/l}$ | 208-387 male<br>149-363 female |
| Alkaline phosphatase | U/l | 30-120 |
| γ-glutamyl transferase | U/l | 0-55 male<br>0-35 female |

CV: cardio-vascular risk

**Table 2.**
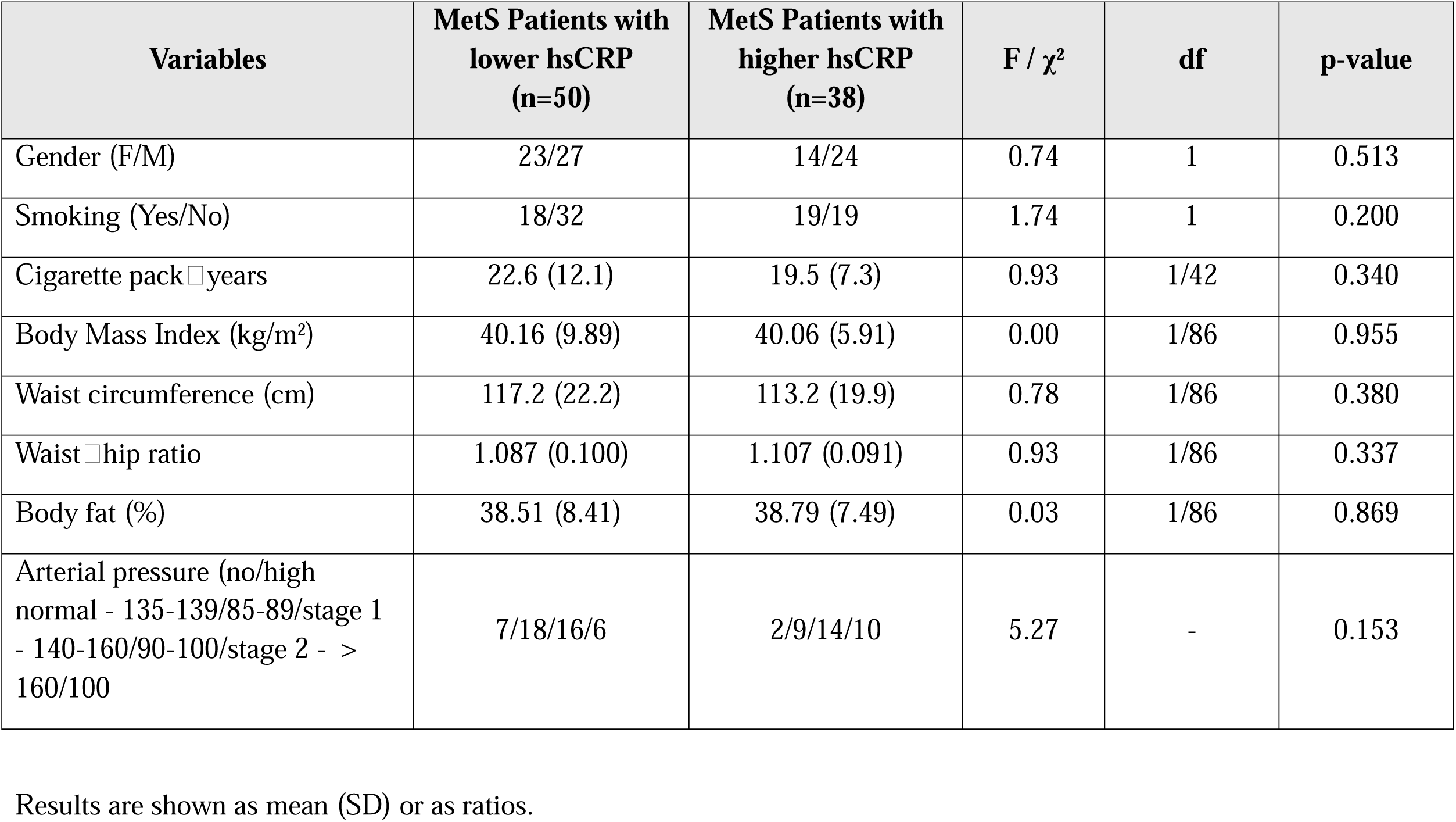
Demographic and clinical data of patients with metabolic syndrome (MetS) and obesity with higher versus lower high sensitivity C-reactive protein (hsCRP) values (13.5 mg/dL cutoff).

### Depression scores and biomarkers in the high versus low-hsCRP Groups

**Table 3** compares the biomarkers among MetS patients dichotomized by hsCRP levels, utilizing a cutoff of 13.5 mg/L. No significant differences were found between the groups regarding both the VZDRtotal and VZDR6 scores. Moreover, there were no differences in serum triglycerides, total cholesterol, HDL, LDL, Castelli Risk Index 1, ApoB/ApoA1 ratio, LAR ratio, apolipoprotein B, apolipoprotein A1, fasting blood glucose, HbA1c, immunoreactive insulin, HOMA-IR, TyG index, leptin, adiponectin, visfatin, IL-6, or TNF-α between both hsCRP groups (even without p-correction). Nevertheless, patients allocated to the high-hsCRP group demonstrated markedly elevated levels of platelet counts, MPV, WBC count, uric acid, ALP, and γ-GT in comparison to individuals with lower hsCRP levels. These differences remained significant after FDR p-correction (at p=0.0034). As a consequence, we have investigated whether one validated PC could be extracted from the hsCRP, platelet counts, MPV, WBC count, uric acid, ALP, and γ-GT data.

**Table 3.** Biomarkers in metabolic syndrome (MetS) patients with higher versus lower high sensitivity C-reactive protein (hsCRP) values (13.5 mg/dL cutoff).

| Variables | MetS Patients with lower<br>hsCRP<br>(n=50) | MetS Patients with<br>higer hsCRP<br>(n=38) | F | df | p-value |
| --- | --- | --- | --- | --- | --- |
| hsCRP (mg/L) | 5.06 (2.56) | 19.76 (4.08) | 429.00 | 1/86 | <0.001 |
| VZDR6 | 11.3 (2.2) | 10.3 (2.2) | 3.65 | 1/86 | 0.059 |
| VZDRtotal | 28.3 (5.8) | 27.5 (5.9) | 0.36 | 1/86 | 0.550 |
| Triglycerides (mmol/L) | 2.75 (1.63) | 2.36 (1.16) | 1.55 | 1/86 | 0.216 |
| Total cholesterol (mmol/L) | 5.84 (1.23) | 5.74 (1.04) | 0.15 | 1/86 | 0.699 |
| HDL□C (mmol/L) | 1.15 (0.27) | 1.20 (0.29) | 0.63 | 1/86 | 0.431 |
| LDL□C (mmol/L) | 3.50 (1.15) | 3.59 (0.76) | 0.16 | 1/86 | 0.692 |
| Apo□B (g/L) | 1.50 (0.48) | 1.51 (0.65) | 0.02 | 1/86 | 0.900 |
| Apo□A1 (g/L) | 1.39 (0.39) | 1.48 (0.41) | 0.96 | 1/86 | 0.329 |
| Castelli risk index 1 | 5.27 (1.49) | 4.96 (1.12) | 1.21 | 1/86 | 0.275 |
| ApoB/ApoA1 | 1.21 (0.63) | 1.19 (0.84) | 0.10 | 1/86 | 0.922 |
| LAR index (LDL/ApoB) | 2.56 (1.14) | 2.79 (1.20) | 0.87 | 1/86 | 0.353 |
| Fasting blood glucose (mmol/L) | 5.84 (1.38) | 6.03 (1.13) | 0.46 | 1/86 | 0.501 |
| HbA1c (%) | 5.87 (0.64) | 5.67 (0.77) | 1.81 | 1/86 | 0.182 |
| Immunoreactive insulin (μIU/mL) | 23.79 (14.16) | 23.05 (17.61) | 0.05 | 1/86 | 0.826 |
| HOMA-IR | 6.41 (4.53) | 6.58 (6.63) | 0.02 | 1/86 | 0.890 |
| TyG index | 0.46 (0.30) | 0.39 (0.22) | 1.60 | 1/86 | 0.209 |
| Leptin (ng/mL) | 64.94 (53.97) | 68.20 (53.79) | 0.08 | 1/86 | 0.779 |
| Adiponectin (μg/mL) | 7.76 (3.88) | 7.98 (3.01) | 0.09 | 1/86 | 0.771 |
| Adiponectin/leptin ratio* | 1.33 (7.61) | 0.39 (1.11) | 0.10 | 1/86 | 0.920 |
| Visfatin (ng/mL) | 5.74 (2.99) | 6.06 (2.52) | 0.30 | 1/86 | 0.587 |
| IL-6 (pg/mL) | 45.05 (92.98) | 53.17 (86.51) | 0.17 | 1/86 | 0.677 |
| TNF-α (pg/mL) | 14.29 (27.18) | 19.11 (10.86) | 1.07 | 1/86 | 0.305 |
| Platelets ($\times 10^9$/L) | 268.46 (80.78) | 394.63 (59.85) | 65.36 | 1/86 | <0.001 |
| MPV (fL) | 9.50 (2.97) | 14.42 (2.97) | 59.18 | 1/86 | <0.001 |
| Leukocytes ($\times 10^9$/L) | 6.68 (2.82) | 10.49 (2.18) | 46.76 | 1/85 | <0.001 |
| Uric acid (μmol/L) | 322.60 (85.21) | 422.74 (41.30) | 44.45 | 1/86 | <0.001 |
| ALP (U/L) | 177.96 (86.69) | 266.74 (68.47) | 27.02 | 1/86 | <0.001 |
| γ-GT (U/L) | 59.48 (22.19) | 82.32 (23.01) | 22.14 | 1/86 | <0.001 |
| LIPUR index (z score) | -0.62 (0.76) | 0.84 (0.60) | 93.35 | 1/85 | <0.001 |

\*Processed in Log transformation; hsCRP: high-sensitivity C-reactive protein, HDL-C: high-density lipoprotein cholesterol, LDL-C: low-density lipoprotein cholesterol, Apo: Apolipoprotein, HbA1c: glycated hemoglobin, HOMA-IR: Homeostatic Model Assessment for Insulin Resistance, TG: Triglycerides, IL: interleukin, TNF: tumor necrosis factor- $\alpha$ , MPV: Mean platelet volume, WBCs: white blood cells, ALP: Alkaline phosphatase, γ-GT: Gamma-glutamyl transferase, LIPUR index: principal component extracted from hsCRP, platelets, MPV, WBCs, Uric acid, ALP, and γ-GT.

### Results of PCA

As shown in **Table 4**, PCA produced one PC1 that explained 65.67% of the total variance in the hsCRP, platelet count, MPV, WBC count, uric acid, ALP, and γ-GT data. All included variables showed factor loadings exceeding 0.70, and the component displayed satisfactory internal consistency, as evidenced by an adequate Cronbach’s alpha value. This PC score was designated as the LIPUR index, reflecting its integration of biomarkers representing liver dysfunction (GGT, ALP), immune activation (hsCRP and leukocytes), platelet activity (platelet count and MPV), and uric acid metabolism.

**Table 4.** Results of Principal Component Analyses (PCA) performed on the biomarkers that are increased in patients with increased high sensitivity C-reactive protein values.

| Variables | Loadings |
| --- | --- |
| hsCRP | 0.705 |
| Platelets | 0.866 |
| MPV | 0.863 |
| WBCs | 0.842 |
| UA | 0.786 |
| ALP | 0.778 |
| γ-GT | 0.820 |
| KMO | 0.925 |
| $\chi^2$ | 470.25 |
| df | 21 |
| p | <0.001 |
| Variance explained | 65.67% |
| Cronbach's alpha | 0.92 |

Abbreviations: KMO, Kaiser-Meyer-Olkin measure of sampling adequacy; $\chi^2$ , Bartlett's test; df, degree of freedom; p, p-value; VE, variance explained; MPV: Mean platelet volume, UA: Uric acid, ALP: Alkaline phosphatase, γ-GT: Gamma-glutamyl transferase.

Consequently, we computed the first PC score and displayed its distribution (see Figure 3). This distribution was clearly bimodal. In addition, we performed a two-step cluster analysis using hsCRP, platelet counts, MPV, WBC count, uric acid, ALP, and γ-GT data as variables. This cluster analysis detected two different clusters of patients which were separated with a silhouette measure of cohesion and separation of 0.7 (more than adequate). This separation into those with high (n=41) versus low (n=46) LIPUR indices coincides with a threshold value of 0.0 applied to the LIPUR index. The latter was significantly higher in MetS patients with high hsCRP versus those with low hsCRP (see Table 3).

**Figure 3.**
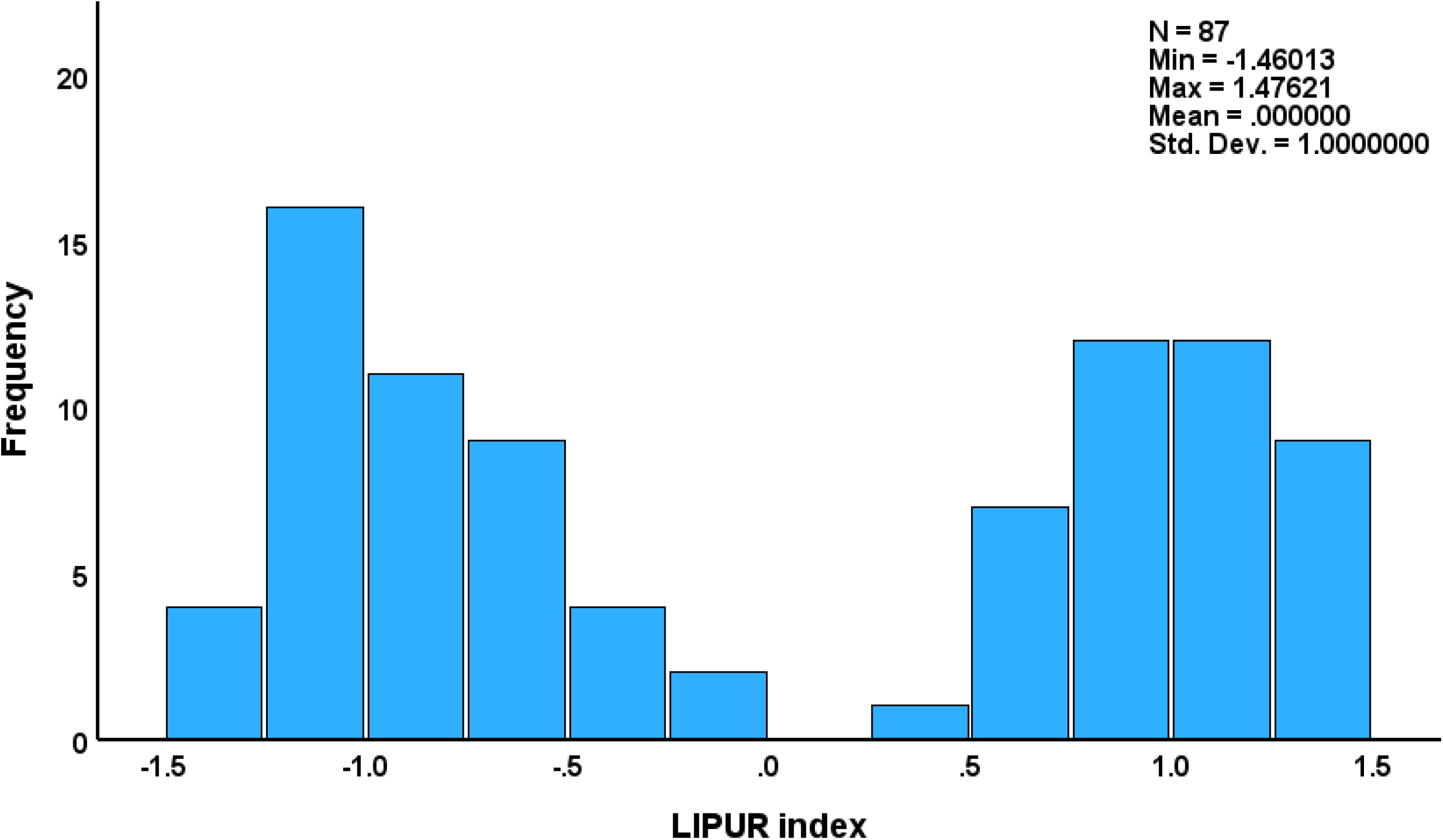
Distribution of the LIPUR index in patients with metabolic syndrome and obesity. The LIPUR index is computed as the first principal component extracted from liver tests, immune and platelet biomarkers and uric acid.

As a consequence, we have computed correlations between severity of depression and the biomarkers in all patients combined and in addition in the two distinct MetS subgroups. The latter distinction is important as there may be qualitative differences in metabolic pathways between both groups which could affect those associations.

### Correlations depression scores and biomarkers

**Table 5** displays the results of multivariate regression analyses examining the relationship between both VZDR scores and the biomarkers in all subjects and in the cluster with low and high LIPUR index, while allowing for the effects of age, sex, BMI, waist-hip ratio, fat mass (%), and smoking status.

**Table 5.** Results of multiple regression analyses with the von Zerssen Depression Severity Rating (VZDR) scores as dependent variables and immune-metabolic biomarkers as explanatory variables.

| Dependent Variables | Explanatory Variables | Coefficients of input variables |  |  | Model statistics |  |  |  |
| --- | --- | --- | --- | --- | --- | --- | --- | --- |
| | | $\beta$ | t | p | R<sup>2</sup> | F | df | p |
| #1a. VZDRtotal in all subjects | Model |  |  |  | 0.198 | 5.07 | 4/82 | <0.001 |
|  | IL-6 | 0.208 | 2.098 | 0.039 |  |  |  |  |
|  | MPV | -0.461 | -3.39 | 0.001 |  |  |  |  |
|  | Leukocytes | 0.350 | 2.57 | 0.012 |  |  |  |  |
|  | TyG | 0.224 | 2.26 | 0.027 |  |  |  |  |
| #1b. VZDR6 in all subjects | Model |  |  |  | 0.276 | 6.16 | 5/81 | <0.001 |
|  | MPV | -0.503 | -3.87 | <0.001 |  |  |  |  |
|  | IL-6 | 0.188 | 1.99 | 0.050 |  |  |  |  |
|  | WBCs | 0.343 | 2.63 | 0.010 |  |  |  |  |
|  | TyG index | 0.293 | 3.05 | 0.003 |  |  |  |  |
|  | Visfatin | -0.216 | -2.25 | 0.027 |  |  |  |  |
| #2a. VZDRtotal in subjects with low LIPUR index | Model |  |  |  | 0.100 | 4.75 | 2/43 | 0.033 |
|  | LDL cholesterol | 0.316 | 2.21 | 0.033 |  |  |  |  |
| #2b. VZDR6 in subjects with low LIPUR index | Model |  |  |  | 0.291 | 8.80 | 2/43 | <0.001 |
|  | Visfatin | -0.452 | -3.51 | 0.001 |  |  |  |  |
|  | ApoB | 0.340 | 2.63 | 0.012 |  |  |  |  |
| #3a. VZDRtotal in subjects with high LIPUR index | Model |  |  |  | 0.142 | 6.45 | 1/39 | 0.015 |
|  | Adiponectin | -0.377 | -2.54 | 0.015 |  |  |  |  |
| #3b. VZDR6 in subjects with high LIPUR index | Model |  |  |  | 0.230 | 5.69 | 2/38 | 0.007 |
|  | MPV | -0.395 | -2.755 | 0.009 |  |  |  |  |
|  | TyG index | 0.328 | 2.285 | 0.028 |  |  |  |  |
| #4 LIPUR index in subjects with the high LIPUR cluster | Model |  |  |  | 0.182 | 3.00 | 1/39 | 0.007 |
|  | IL-6 | 0.382 | 2.874 | 0.007 |  |  |  |  |

LIPUR index: integrated index of Liver + Uric acid + Platelet + Inflammatory biomarkers, MPV: Mean platelet volume, WBCs: White blood cells count, TG: Triglyceride, Apo: Apolipoprotein, TyG: triglyceride x fasting blood glucose index.

In the total study sample (regression #1a and #1b), 19.8% of the variance in the VZDRtotal score was explained by the regression on IL-6, TyG and leukocytes (positively) and MPV (inversely). 27.6% of the variance in the VZDR6 score was explained by five significant predictors, namely MPV and visfatin (both inversely associated), and IL-6, WBC count and TyG ratio (positively associated). Figure 4 shows the partial regression of VZDR6 score on the TyG index.

**Figure 4.**
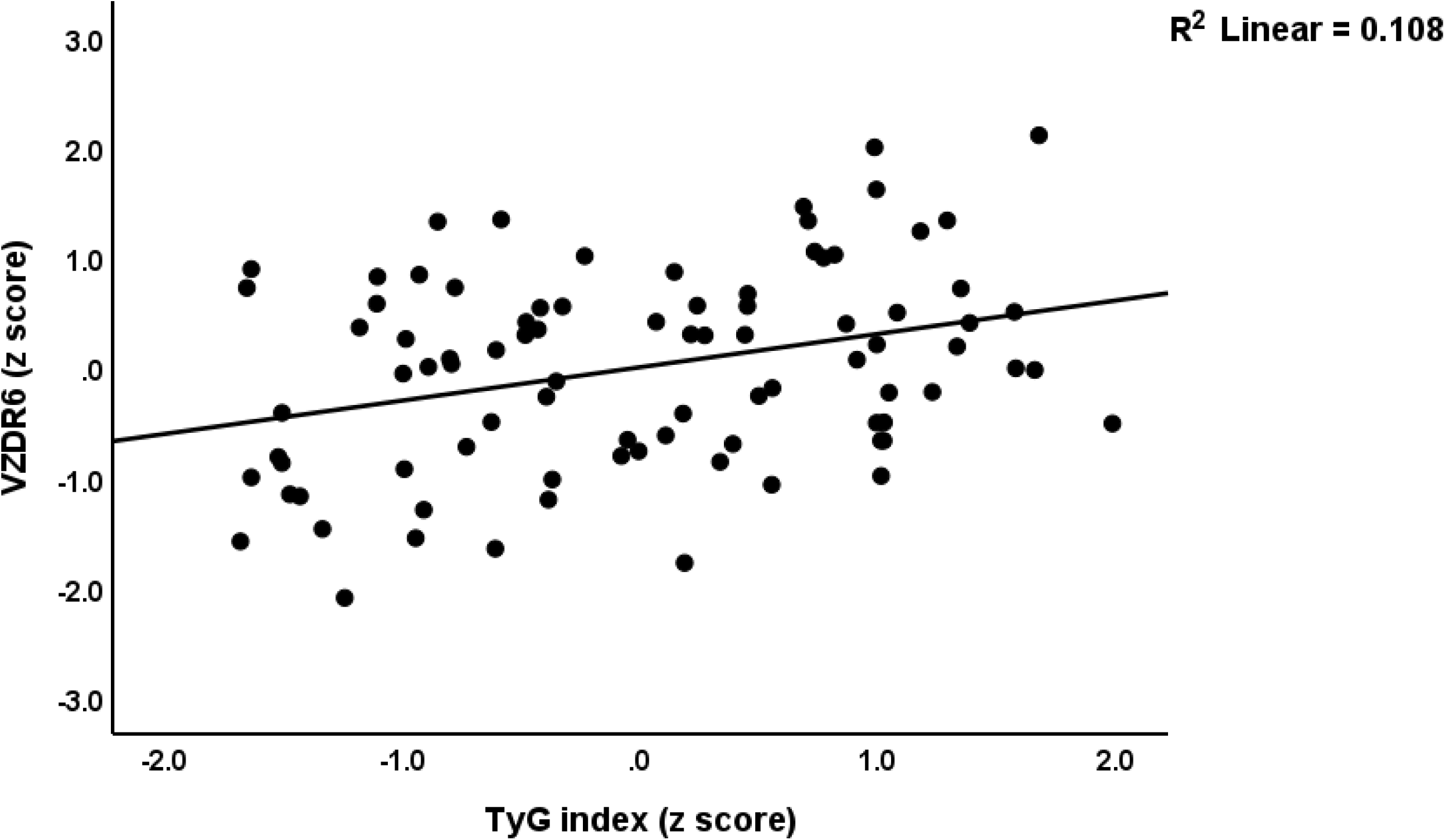
Partial regression of the von Zerssen Depression Rating (VZDR6) scale score on the TyG index (p<0.01)

In the low LIPUR subgroup (regression #2a and #2b), 10.0% of the variance in VZDRtotal score was explained by LDL (positively associated). 29.1% of the variance in the VZDR6 score was accounted for by visfatin (negatively associated) and apolipoprotein B (positively associated). Figures 5 **and 6** show the partial regression plots illustrating the adjusted relationship between the VZDR6 score and visfatin and AopB, respectively. In the high LIPUR subgroup (regressions #3a and #3b), lower adiponectin was associated with the VZDRtotal score explaining 14.2% of the variance. MPV was negatively associated, while the TyG ratio was positively associated with the VZDR6 score. These two variables jointly explained a significant proportion (23%) of the variance in this score. Additional regression analysis (regression #4) indicated that 18.2% of the variance in the LIPUR index in the high LIPUR cluster could significantly be predicted by IL-6.

**Figure 5.**
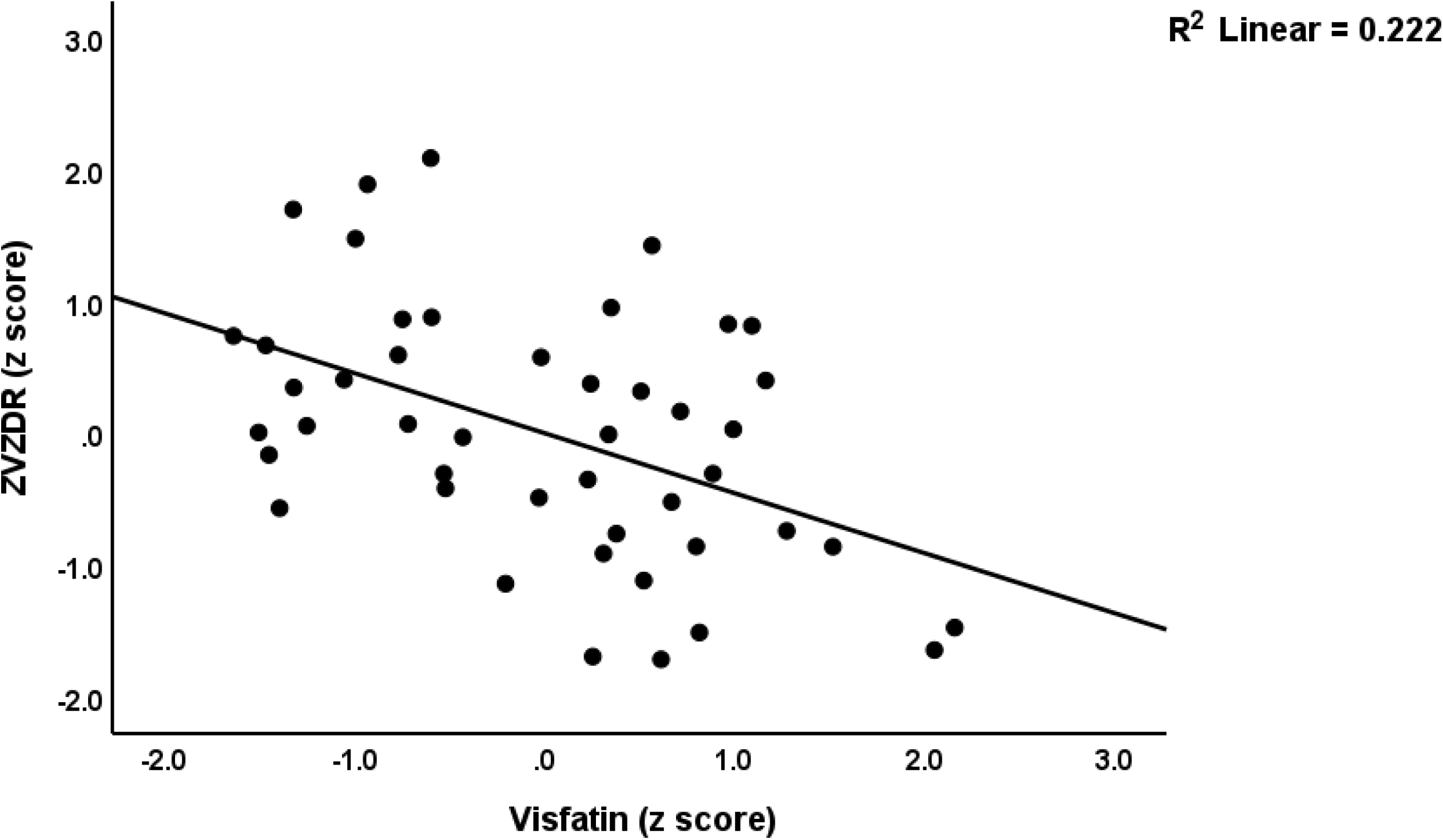
Partial regression plot illustrating the relationship between the subdomain of the von Zerssen Depression Rating (VZDR6) scale sore on serum visfatin (p<0.01)

**Figure 6.**
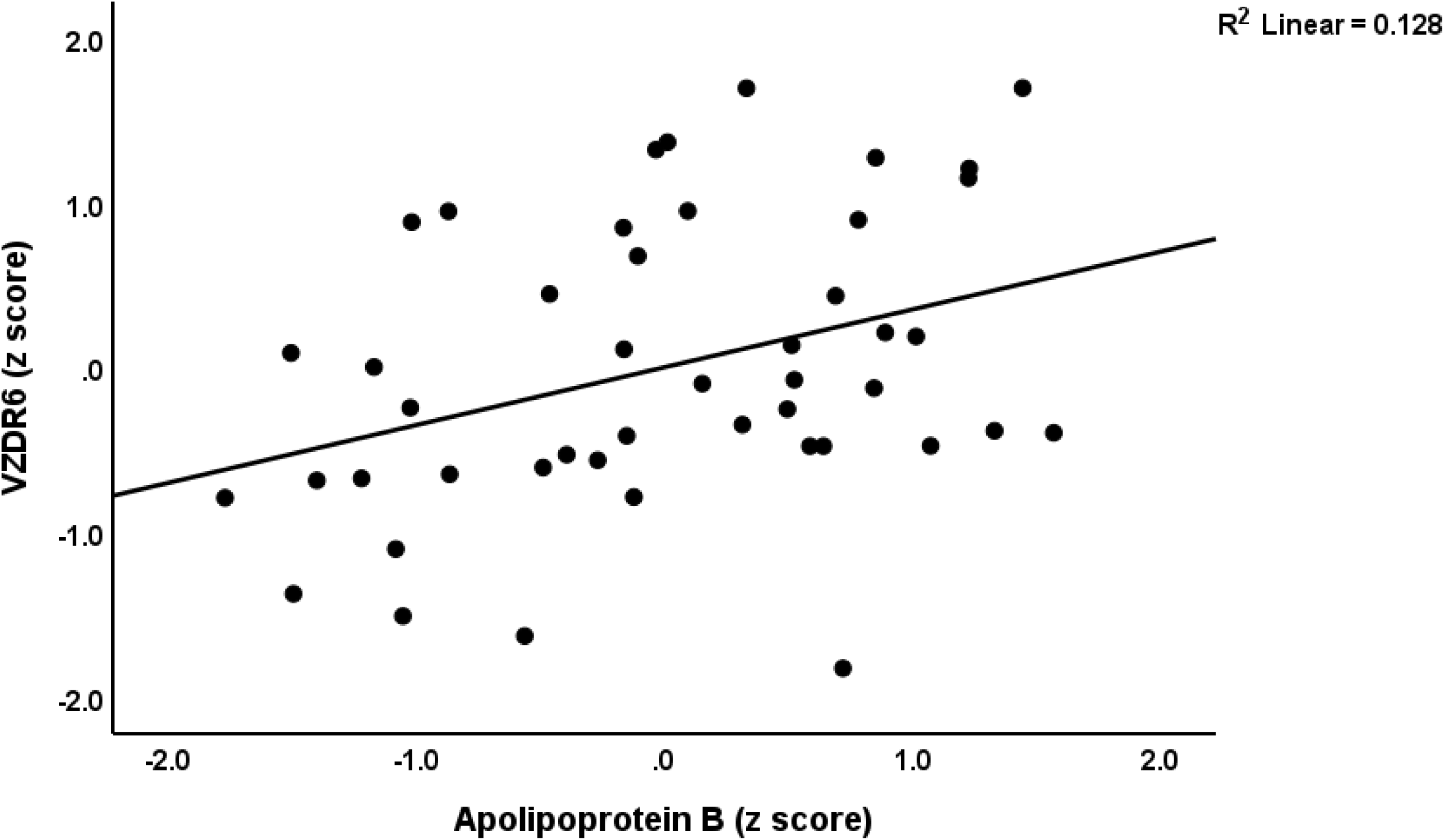
Partial regression plot illustrating the relationship between the subdomain of the von Zerssen Depression Rating (VZDR6) scale sore on serum apolipoprotein B (p<0.01)

### Depression scores and biomarkers in the LIPUR subclasses

There were no significant differences in the VZDR6 (F = 2.00, df = 1/85, p = 0.161) and VZDRtotal (F=0.88, df=1/85, p=0.350) scores between patients with a lower LIPUR index and those with a higher index **Table 6** summarizes the correlation analysis between the LIPUR index and depression severity, immune, and metabolic scores. In both the low and high-LIPUR index clusters, no significant correlations were observed between the LIPUR indices and the depression severity scores.

**Table 6.** Intercorrelation matrix (Pearson’s correlation coefficients) between the LIPUR index and different immune-metabolic biomarkers in both clusters of metabolic syndrome patients.

| Variables | LIPUR index (in the low-LIPUR cluster, n=46) | LIPUR index (in the high-LIPUR cluster, n=41) |
| --- | --- | --- |
| VZDR6 | -0.124 (0.411) | -0.125 (0.438) |
| VZDRtotal | 0.078 (0.604) | -0.063 (0.694) |
| Interleukin-6 | -0.122 (0.417) | 0.386* (0.013) |
| Tumor necrosis factor-α | 0.127 (0.400) | 0.342* (0.029) |
| Fasting blood glucose | 0.177 (0.239) | 0.340* (0.030) |
| Immunoreactive insulin | 0.127 (0.401) | 0.343* (0.028) |
| HOMA-IR | 0.211 (0.160) | 0.341* (0.029) |
| Glycated hemoglobin | 0.248 (0.097) | 0.315* (0.045) |

LIPUR index: principal component extracted from high sensitivity C-reactive protein (hsCRP), platelet number, mean platelet volume, number of leukocytes, uric acid, alkaline phosphatase, gamma-glutamyl transferase, HOMA-IR: Homeostatic Model Assessment for Insulin Resistance, IL: interleukin, TNF: tumor necrosis factor.

In both patient clusters there were no significant correlations between the LIPUR index and any of the atherogenic data. In patients allocated to the high LIPUR cluster, but not in those allocated to the other cluster, we found significant associations between the LIPUR index and both cytokines, and insulin resistance data.

## Discussion

### Depressive symptoms in MetS

The first major finding of this study indicates that the severity of depressive symptoms measured using the VZDR scale scores did not associate with increased hsCRP levels. The findings are consistent with earlier studies that did not demonstrate a reliable association between hsCRP and depressive symptoms (Krogh et al., 2014a, Mommersteeg et al., 2014, Khan et al., 2020).

In contrast to the lack of associations between hsCRP and depressive symptoms, this study observed that severity of depression correlated mainly with immune activation (indicated by increased IL-6 and leukocyte number), and aberrations in platelets (lower MPV), atherogenicity (increased ApoB and LDL), insulin resistance (increased Ty G ratio), and lower visfatin and adiponectin.

IL-6, which induces CRP production, and increased numbers of leukocytes were associated with depressive symptoms, although the effect size of IL-6 was rather small. It is well known that depression is associated with increased serum levels of IL-6 (Maes et al., 1995, Osimo et al., 2020) and with increased numbers of leukocytes (Maes et al., 1993b, Foley et al., 2023). Depression is characterized by dysregulation in neuro-immune–metabolic–oxidative (NIMETOX) pathways (Maes et al., 2025). Jirakran et al. (2025) recently reported that lipid disturbances in MDD, characterized by reduced reverse cholesterol transport and heightened atherogenicity, cannot be effectively assessed in the presence of MetS (Jirakran et al., 2025b). Recent meta-analyses found that mood disorders, including MDD and bipolar disorder, are associated with increased atherogenic indices such as the Castelli Risk Index-2, the Atherogenic Index of Plasma (Almulla et al., 2023, Jirakran et al., 2025a). Recently, Maes et al. (2024) introduced the concept of an “atherommune-phenome” in outpatient MDD patients, highlighting the critical role of heightened atherogenicity linked to immune-mediated neurotoxicity in depressive pathophysiology. The alteration between MetS and MDD aggravates cytokines production and worsening symptoms (Maes et al., 2024b). Moreover, increased insulin resistance indices contribute to the severity of depression in patients with major depression (Maes et al., 2025).

There is now some evidence that depression is accompanied by lower serum levels of adiponectin (Hu et al., 2015, Islam et al., 2022). This hormone, synthesized by adipose tissue, exhibits antioxidant, anti-inflammatory, and anti-atherogenic properties that promote metabolic and cardiovascular health (Ohashi et al., 2012). Experimental studies show that exogenous adiponectin produces antidepressant-like effects in animal models of stress-induced depression, indicating a potential role in mood regulation (Liu et al., 2012). Additionally, baseline adiponectin levels affect the effectiveness of different antidepressant combination therapies in MDD (Furman et al., 2018). Adiponectin is increasingly recognized as a potential therapeutic target in central nervous system disorders, including depression, owing to its neuroprotective and mood-regulating properties (Bloemer et al., 2018).

Visfatin is a proinflammatory adipokine that is secreted by visceral fat and is involved in insulin signaling and resistance (Alghasham and Barakat, 2008, Stofkova, 2010). Higher visfatin levels may be detected in depression (Akinlade et al., 2020) and the remission phase of inflammatory disease like rheumatoid arthritis or inflammatory bowel disease (Lee and Bae, 2018, Saadoun et al., 2021). Nevertheless, increased insulin levels might lower visfatin production (Kowalska et al., 2013). Importantly, visfatin (and other adipokines) has neuroprotective effects via enhancing cellular energy metabolism, and it may have glucose-lowering and insulin-enhancing effects (Fukuhara et al., 2005, Erfani et al., 2015, Lee et al., 2019). Thus, while increased visfatin is a marker of obesity and MetS that may aggravate lipid aberrations, atherosclerosis and type 2 diabetes mellitus, lower levels may reduce neuroprotection, thereby possibly aggravating the pathophysiology of depression.

Lower MPV values have been found in some, but not all studies, in depression particularly following treatment (ÖZTüRK et al., 2019). One explanation is that IL-6 and glucocorticoid levels, which are both increased in depression (Maes et al., 1993a) could have caused lower MPV values (Clarke et al., 1996). Nevertheless, in the current study, many MetS patients (those belonging to the high LIPUR cluster) display high MPV values. As such, the lowered MPV levels in our MetS patients may be regarded as a secondary phenomenon not directly related to the pathophysiology of depression.

Overall, our findings indicate that increasing severity of depression in our drug-free MetS patients is an indicant of immune activation, atherogenicity, insulin resistance and lowered antioxidant defenses. Previous studies have repeatedly emphasized the association between particular depressive symptom profiles and components of MetS. Suttajit and Pilakanta revealed that symptoms including depressed mood, insomnia, and psychomotor retardation are significantly linked to central obesity and hyperglycemia, indicating that specific depressive phenotypes may demonstrate stronger metabolic associations (Suttajit and Pilakanta, 2013). Chirinos et al. (2017) identified four distinct profiles of depressive symptoms among healthy mid-life adults in the United States, each linked to different cardiometabolic outcomes (Chirinos et al., 2017).

Therefore, the issue remains: why is hsCRP not a biomarker of depression in MetS, whereas IL-6, which induces CRP production, is associated with depression? The following section will address this matter.

### Distinct subgroups of MetS patients

The second major finding of this study is that MetS in obese patients comprises distinct subclasses. Thus, PCA and cluster analysis revealed two biologically significant clusters based on higher and interrelated levels of hsCRP, uric acid, ALP, γ-GT, WBC counts, MPV, and platelet counts.

Mild to moderate inflammation frequently results in an increase in the number of platelets and an increase in MPV (Thomas and Storey, 2015, Korniluk et al., 2019). Thrombocytosis may be induced by IL-6 through an increase in thrombopoietin production (Kaser et al., 2001). Furthermore, megakaryopoiesis induced by IL-6 or TNF-α may also result in an increase in MPV, which is characterized by activated, larger, and younger platelets (Gardner and Bessman, 1983, Dan et al., 2009). Metabolic or hepatic inflammation may be indicated by elevated ALP and γ-GT levels. For instance, IL-6 may elicit both enzymes by promoting the acute phase response and activating hepatocytes (Toth et al., 2004, Schmidt-Arras and Rose-John, 2016). The production of uric acid can also be induced by IL-6 through stimulation of xanthine oxidase (Pfeffer et al., 1994), and impaired renal excretion of uric acid (Tsutani et al., 2000, Zhang et al., 2024). Intertwined associations between these LIPUR pathways and pro-inflammatory cytokines (IL-6, TNF-α) and insulin resistance indices (HOMA-IR and TyG indices and HbA1c) further characterize this particular cohort of MetS patients. As a result, the cluster of MetS patients with high LIPUR indices is distinguished by a substantially increased inflammatory load, which is induced by pro-inflammatory cytokines, as well as elevated liver enzymes, uric acid levels, and heightened immune and platelet activation.

It is important to emphasize that the subgroup with a lower LIPUR index also demonstrates substantial immune-metabolic abnormalities, specifically elevated hsCRP and IL-6 levels, HOMA-IR, TyG, Castelli risk index 1, and liver tests in comparison to the normal values (refer to Table 1). However, hsCRP is a biomarker of a specific immune-metabolic subtype of MetS, identified as the high-LIPUR subclass. This subclass is not associated with depression. Depression is more closely linked to other immune-metabolic pathways, including insulin resistance and atherogenicity, as well as diminished antioxidant defenses.

The results of the current study emphasize that, in MetS, elevated serum hsCRP levels cannot be used as a biomarker of inflammation in association with depression. Indeed, increased hsCRP is a marker of a specific subgroup of MetS, characterized by specific immune-metabolic LIPUR alterations. In depression, approximately 50% of the variance in hsCRP can be attributed to factors that are not associated with mood disorders, such as an elevated BMI and advanced age (Moraes et al., 2017). Based on abdominal adiposity, hyperglycemia, and BMI, Junqueira et al. demonstrated that CRP is a reliable indicator of MetS (Junqueira et al., 2009). Cardiovascular risk is frequently indicated by even minor increases in hsCRP within lower ranges, which may result from conditions such as obesity, subclinical atherosclerosis, or metabolic syndrome (Blaha et al., 2011, Timpson et al., 2011). Given that hsCRP is widely recognized as a marker of the metabolic-inflammatory burden associated with MetS (Mirhafez et al., 2016, Maes, 2025), our results substantiate the assumption that elevated hsCRP primarily indicates metabolic disturbances that are inherent to MetS, and in particular for one MetS subtype (the high LIPUR subgroup), rather than serving as a specific marker for depression.

## Limitations

It is essential to acknowledge some limitations while interpreting the current findings. The existence of two different MetS subgroups with respect to the LIPUR index deserves replication in other countries and cultures and in a cohort of MetS patients without obesity. The present study would be even more interesting if we could examine additional cytokines, chemokines, and growth factors alongside oxidative and nitrosative stress biomarkers that are associated with depressive symptoms and MetS (de Melo et al., 2017).

## Conclusion

This study illustrates that hsCRP is insufficient as a biomarker for depressive symptoms in patients with MetS, even when individuals exhibit a substantial inflammatory burden. The depressive symptoms in MetS are predominantly associated with atherogenicity (increased ApoB and LDL), insulin resistance (increased TyG index), lower visfatin and adiponectin levels, but also with immune activation indicators (IL-6 and leukocyte number), and platelet aberrations (lower MPV). Elevated hsCRP levels in MetS are significantly correlated with LIPUR indicators indicative of liver dysfunction, uric acid, platelet activation, and immunological activation, a phenomenon seemingly influenced by heightened production of IL-6 and TNF-α. There are no associations between depressive symptoms and either hsCRP or both MetS phenotypes. These results again indicate that combining individuals with and without MetS/obesity to examine the interrelations between depression, hsCRP and metabolic biomarkers is obsolete.

The identification of two distinct LIPUR subgroups emphasizes the heterogeneity of MetS and reinforces the need for specific assessment of concomitant symptoms and treatment modalities. Individuals in the lower-LIPUR subgroup may necessitate interventions aimed at lifestyle adjustments and pharmacological therapies to treat atherogenicity and insulin sensitivity and improve metabolic health. Patients with the high-LIPUR phenotype may benefit from supplementary targeted therapy designed to mitigate hepatic dysfunction, elevated uric acid synthesis, the activation of platelets, and increased IL-6 and TNF-α production.

## Data Availability

The corresponding author (MM) will provide access to the dataset supporting this study upon receipt of a valid request and the completion of a thorough data review.

## Acknowledgements

Not Applicable

## Ethical approval and consent to participate

The study was approved by the Research Ethics Committee of the Medical University – Plovdiv (Protocol No. 22; 31 December 2023). Written informed consent was obtained from the participants.

## Declaration of interest

No competing of interest to be declared by authors.

## Author’s contributions

All authors contributed equally to this research and approved the final version of the paper.

## Notes

### Competing Interest Statement

The authors have declared no competing interest.

### Funding Statement

None

### Author Declarations

The study was approved by the Research Ethics Committee of the Medical University of Plovdiv (Protocol No. 22; 31 December 2023). Written informed consent was obtained from the participants.

